# Effectiveness of resting transthoracic echocardiography at detecting significant coronary artery disease in adults

**DOI:** 10.1101/2023.11.01.23297950

**Authors:** Michael D. Woods, Jess Hatfield, Kendall Hammonds, Alex Pham, Jose Exaire, Timothy Mixon, Vinh Nguyen, Christopher Chiles, Robert J. Widmer

## Abstract

**Background:** Detection of regional wall motion abnormalities (RWMA) on TTE is a commonly used to correlate for coronary artery disease (CAD) and often prompts a further workup, including cardiac computed tomography (CT) or cardiac catheterization. However, RWMAs do not consistently predict obstructive CAD. This study investigates the accuracy and reliability of the presence of RWMA on TTE at detecting significant CAD (≥ 70 % vessel stenosis). **Methods:** A retrospective chart review was conducted of adults seen by the Baylor Scott & White Temple echocardiography laboratory who received a resting TTE followed by cardiac catheterization within 30 days over a 4-year period. Exclusion criteria included catheterization without coronary angiography and prior history of CAD, percutaneous coronary intervention (PCI), or coronary artery bypass graft (CABG). We analyzed RWMA on TTE and atherosclerotic CAD on cardiac catheterization to assess for correlation. **Results:** 435 patients were included in the study and 198 patients received ultrasound enhancing agent (UEA). The sensitivity and specificity of RWMA on TTE for detecting CAD in adults was 49.5 % and 78.8%, respectively. The positive and negative likelihood ratios were 2.33 and 0.641, respectively. The use of UEA made no significant difference in the sensitivity or specificity. **Discussion:** Our results show that the presence of RWMA on TTE has a high ability to rule in CAD but the absence of RWMA displays a much lower ability to rule out CAD than previously reported. Our results also show UEA did not enhance or detract this relationship. Clinicians should be aware that the presence of RWMA on resting TTE has a high association with obstructive CAD but the absence of RWMA does not sufficiently exclude CAD.

**Clinical Perspective:** Current guidelines support the use of resting TTE in suspected occlusive coronary artery disease when ECG, biomarkers, and patient history are insufficient to warrant cardiac catheterization. Our study demonstrated the presence of RWMA on resting TTE has a high association with obstructive CAD but the absence of RWMA does not sufficiently exclude CAD. While a positive TTE provides good evidence for additional workup of CAD, practicing clinicians should carefully weigh their plan in the event of a negative TTE prior to ordering the test to determine if the test is a necessary diagnostic step for their patient. If the clinician would decide to continue with a workup for CAD despite a negative resting TTE, the clinician may consider skipping the TTE and moving straight to their further workup to increase the economic value of care provided.

## Introduction

Heart disease is the leading cause of premature mortality in the United States and coronary artery disease (CAD) has remained the leading cause of death in the United States for the last three decades.(1) Although cardiac catheterization for coronary angiogram is the gold standard for diagnosing CAD, its invasive nature renders it impractical for ubiquitous use and necessitates appropriate screening of patients. Electrocardiography (ECG) and cardiac biomarkers are employed in suspected acute coronary syndromes (ACS); however, they are supportive but not always diagnostic. Non-invasive cardiac imaging techniques, such as computed tomography (CT), cardiac magnetic resonance (CMR), positron emission computed tomography (PET), and transthoracic echocardiography (TTE), are further capable of elucidating whether a patient has coronary artery disease.(2) Among these techniques, TTE is indicated in evaluation of suspected myocardial infarction (MI) when patient history, ECG, and biomarkers are unable to sufficiently characterize a patient due to its rapidity, availability, cost-effectiveness, and its lack of ionizing radiation.(3)

Although stress echocardiography has been established as a useful tool for the diagnosis of chronic CAD, it is contraindicated in high-risk ACS, decompensated heart failure (HF), or patients with significant arrythmias. In these acute presentations of CAD, resting TTE is typically utilized and may provide more definitive information than assumed. TTE may detect evidence of myocardial dysfunction in the form of regional wall motion abnormalities (RWMA), the presence of which may even precede ECG changes.(4)

RWMA abnormalities are not specific to CAD and several previous studies reported a significantly higher sensitivity than specificity for TTE to identify CAD(5–7), and there is some controversy regarding the efficacy of using TTE to diagnose lesion/vessel specific CAD.(8,9) These previous studies are limited by restrictive patient populations and/or small sample sizes and establish the need for evaluating the diagnostic usefulness of resting TTE in patients with CAD.

This study evaluates the effectiveness of resting TTE done prior to coronary angiography at detecting significant CAD (≥ 70% vessel occlusion) in adults to better inform how resting TTE results can serve as an adjunct modality to diagnose vessel-specific CAD.

## Materials and Methods

### Study design and patient selection

All patients admitted to a large tertiary care facility in Temple, TX or seen outpatient by the Baylor Scott and White Temple Cardiology group between January 1, 2019, and December 31, 2022, who satisfied the following criteria were included in this retrospective study: (1) ≥ 18 years of age, (2) received a cardiac catheterization, and (3) received a resting TTE within 30 days prior to the catheterization. Exclusion criteria included patients who did not receive a coronary angiogram during their catheterization or had a prior history of CAD, percutaneous coronary intervention (PCI), or coronary artery bypass graft (CABG).

A comprehensive electronic medical record review of included participants was conducted to collect the patient’s age, sex, race, ethnicity, body mass index (BMI), and past medical history at the time of the catheterization. Past medical history collected included congenital heart disease, amyloidosis, sarcoidosis, hemochromatosis, peripheral vascular disease (PVD), stroke, chronic kidney disease, chronic liver disease, primary cancers, chemotherapy, chronic obstructive pulmonary disease (COPD), obstructive sleep apnea (OSA), asthma, chronic heart failure, and cardiac rhythm abnormalities. Additionally, TTE and cardiac catheterization reports were reviewed for the information outlined below and the time between when the TTE and catheterization took place.

### *Transthoracic* echocardiography

Transthoracic echocardiography (TTE) was performed using conventional 2- dimensional projection, spectral and color Doppler, and M-mode.(10) TTE reports were reviewed for indication for TTE by ICD-10 (International Classification of Disease-10) code, use of ultrasound enhancing agent (UEA), ejection fraction (EF), and the presence of any valvular abnormalities, systolic dysfunction, diastolic dysfunction, technical difficulties, global hypokinesis, or RWMA. UEA was administered when two or more contiguous segments were suboptimally visualized. RWMA was defined as one or more hypokinetic, dyskinetic, aneurysmal, or akinetic segments of any of the 17 myocardial segments outlined in the American Heart Association guidelines. Global hypokinesis was not considered to be positive for RWMA unless there were discrete, superimposed akinetic segments identified.

### Coronary catheterization and angiography

Coronary angiography reports were reviewed for the diagnostic angiogram findings detailing stenosis of any observed vessel. Significant coronary artery disease (CAD) was defined as ≥ 70% vessel stenosis of one or more of the major coronary arteries and/or their major branches.

### Data analysis

Continuous variables are expressed as the mean and standard deviation and compared with a 2-tailed Student’s t-test with equal or unequal variance as determined by an F-test. Categorical variables are expressed as frequencies and proportions, compared with either χ^2^ or Fisher exact tests, as appropriate, and reported as odds ratios (OR) with a 95% confidence interval (CI). Breslow-Day- Tarone tests were used to assess the homogeneity of odds ratios across a third categorical variable. Multivariate logistic regression analysis was conducted to identify possible confounding variables affecting the relationship between RWMA and CAD, and a receiver operating characteristic (ROC) curve was generated adjusting for identified confounding variables. Sensitivities and specificities are reported with a 95% CI and were compared using χ^2^ tests. Statistical significance was determined by p-values less than 0.05. All statistical analyses were performed in SAS 9.4 (Cary, NC).

## Results

959 patients were identified as candidates. 384 patients were duplicates, 80 patients did not receive coronary angiography, and 60 patients had a prior history of CAD, PCI, or CABG, resulting in 435 patients ultimately being included in this study. The mean age was 62.0 ± 14.2 years old and mean BMI was 30.2 ± 7.2 kg/m^2^. 245 patients were male (56.3%), 336 patients were white (77.2%), and 56 were Hispanic/Latino (12.9%). Additional patient characteristics are shown in Table 1.

**Table 1.**
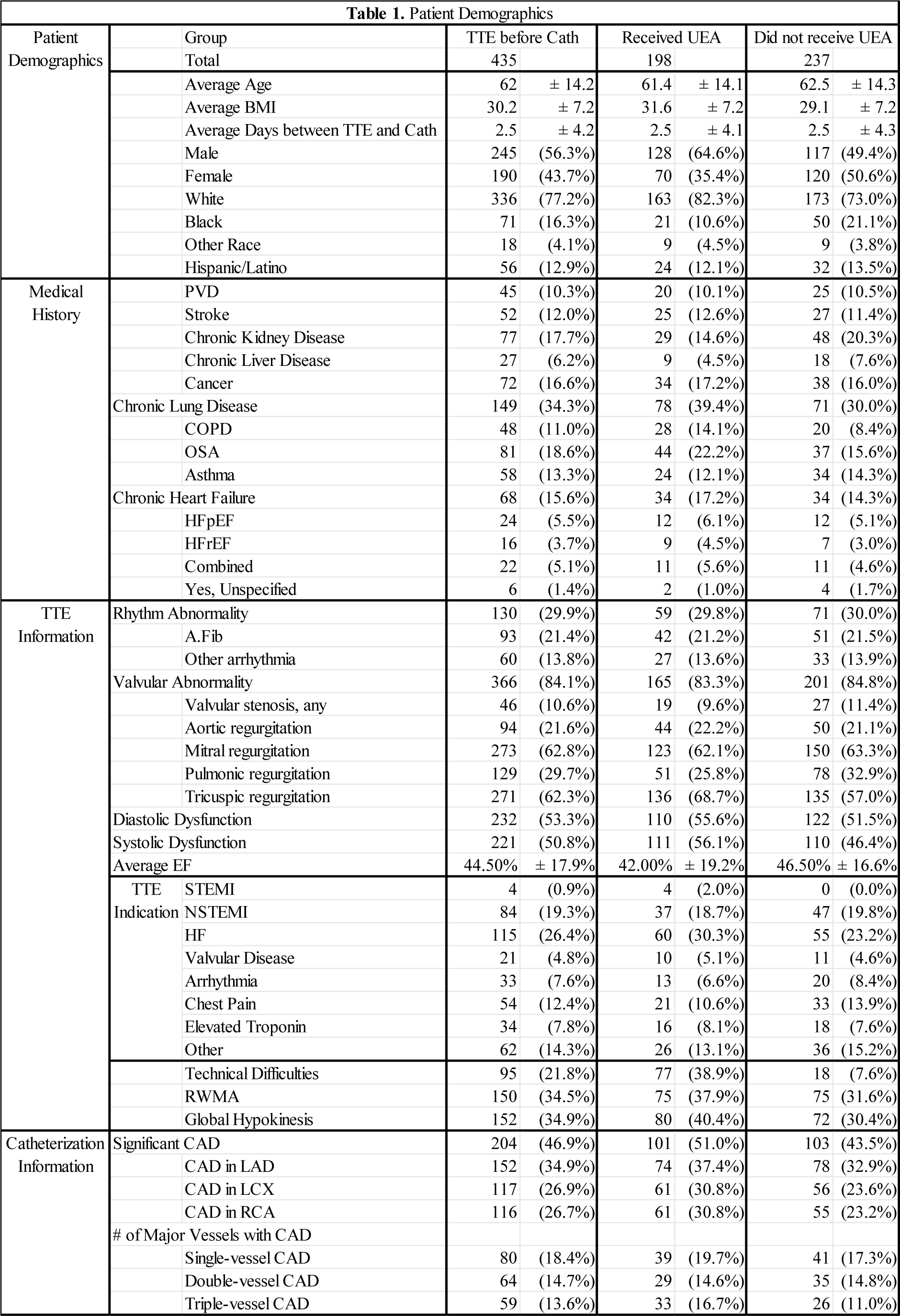
Study population demographics by the presence of coronary artery disease (CAD) with and without regional wall motion abnormalities (RWMA) and use of ultrasound enhancing agent (UEA) on resting transthoracic echocardiography (TTE).

Of the 435 included patients, 204 were found to have CAD, 150 were found to have RWMA, and 101 were found to have both CAD and RWMA. The sensitivity and specificity of RWMA on resting TTE at detecting CAD was 49.5% (95% CI: 48.2 – 50.9%) and 78.8% (95% CI: 69.0 – 88.6%), respectively. The positive and negative likelihood ratios were 2.33 (95% CI: 1.67 - 3.00) and 0.64 (95% CI: 0.54 - 0.74), respectively (Table 2).

**Table 2.**
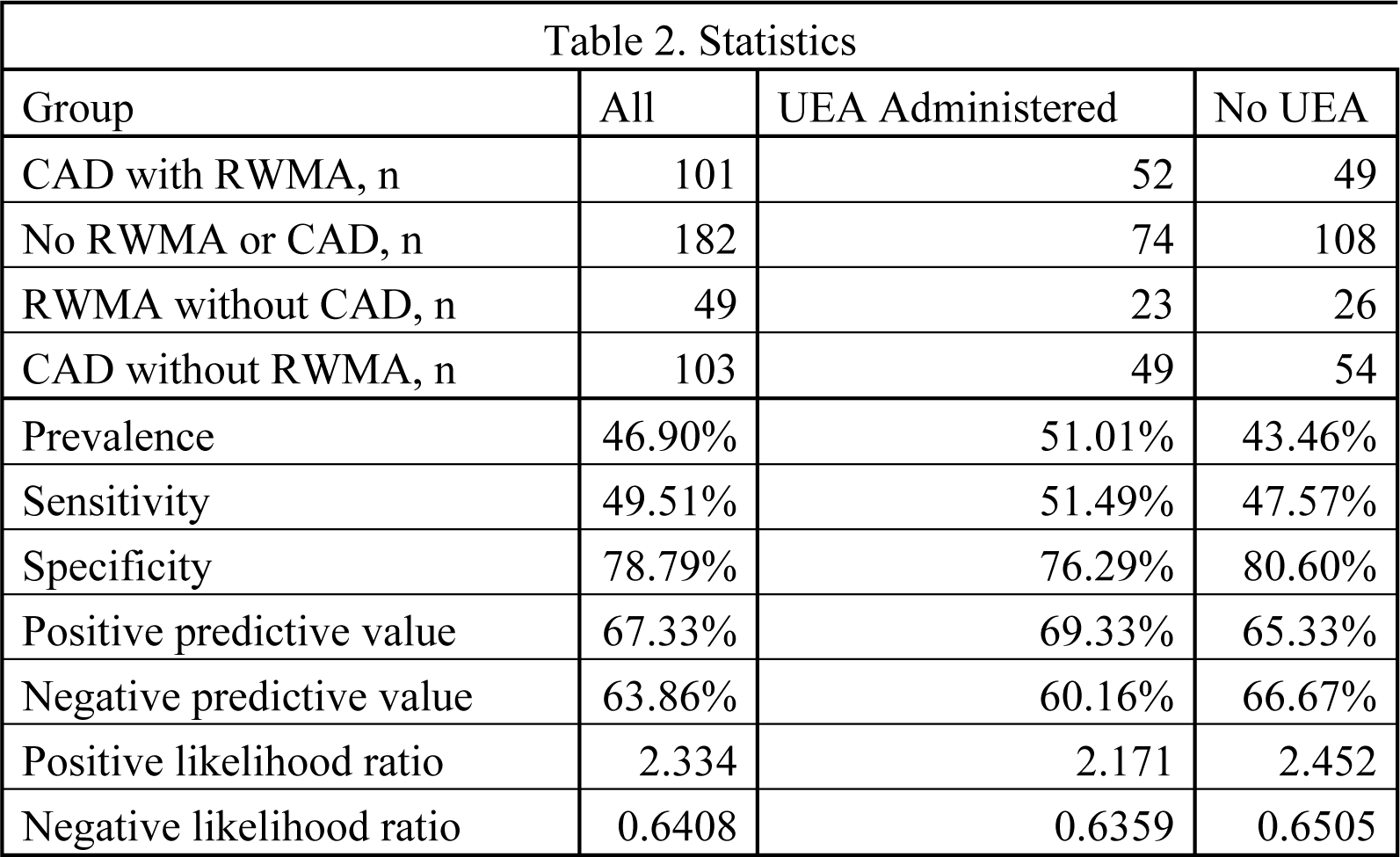
Diagnostic effectiveness of regional wall motion abnormalities (RWMA) on resting transthoracic echocardiography (TTE) at detecting significant coronary artery disease (CAD, ≥ 70% vessel occlusion).

As shown in Figure 1,multivariate logistic regression identified a BMI ≥ 30 kg/m^2^ (OR = 0.64, 95 % CI: 0.42 – 0.972, p = 0.04), the presence of systolic or diastolic dysfunction (OR = 0.56, 95 % CI: 0.35 – 0.89, p = 0.01), and a delay between TTE and cardiac catheterization greater than 2.5 days (OR = 0.57, 95 % CI: 0.36 – 0.90, p = 0.02) independently modified the relationship between RWMA and CAD. A history of COPD, rhythm abnormalities, the presence of a pacemaker or internal cardiac defibrillator, and the administration of UEA were not significant modifiers of the relationship between RWMA and CAD. The area under the curve (AUC) of the ROC curve for RWMA predicting CAD, adjusting for the above significant modifiers, was 0.7136 (Figure 2).

**Figure 1.**
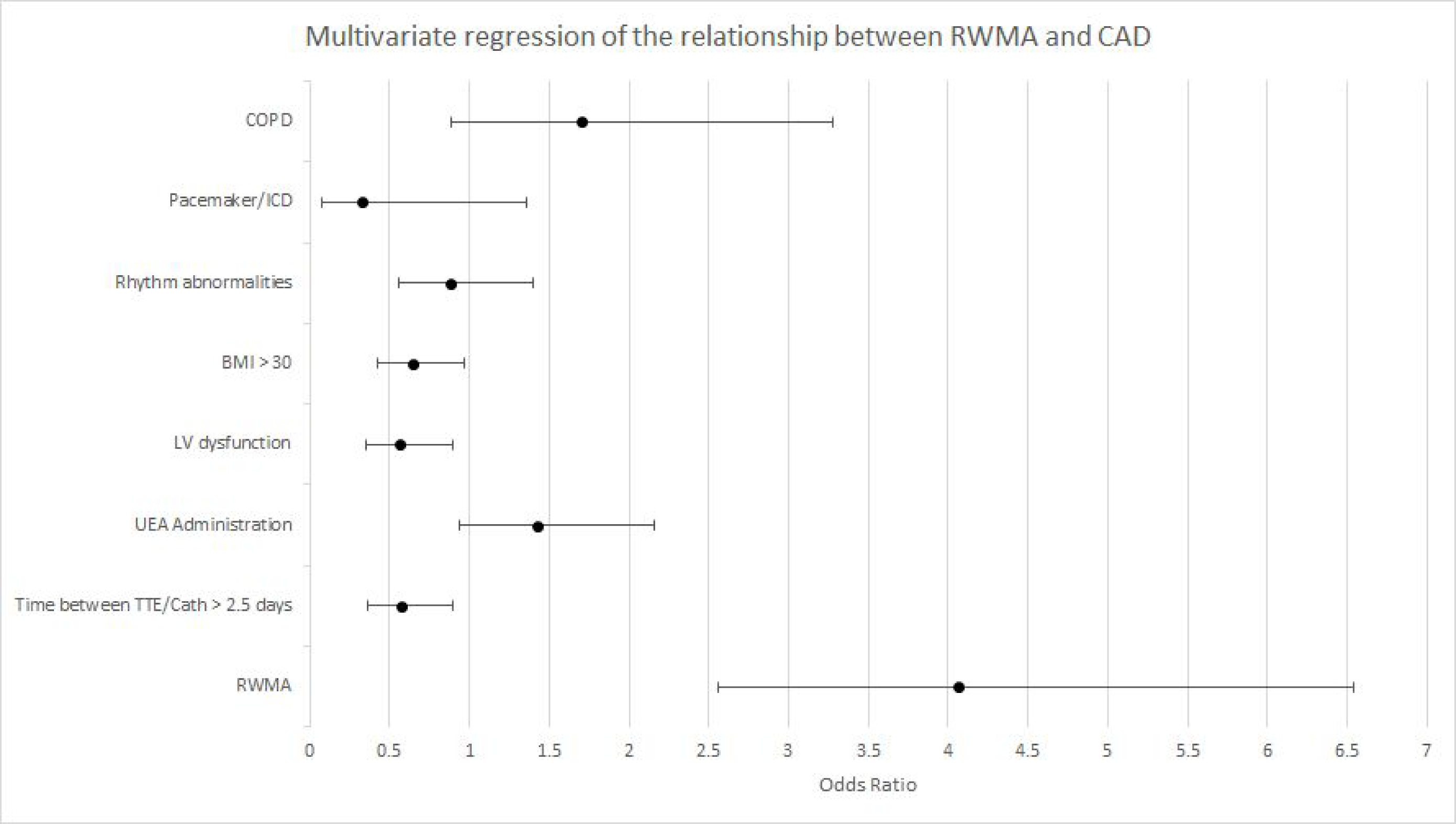
Odds ratio plot of multivariate logistic regression of the relationship between regional motion wall abnormalities (RWMA) on resting transthoracic echocardiography (TTE) and coronary artery disease (CAD) adjusting for potential confounding variables.

**Figure 2.**
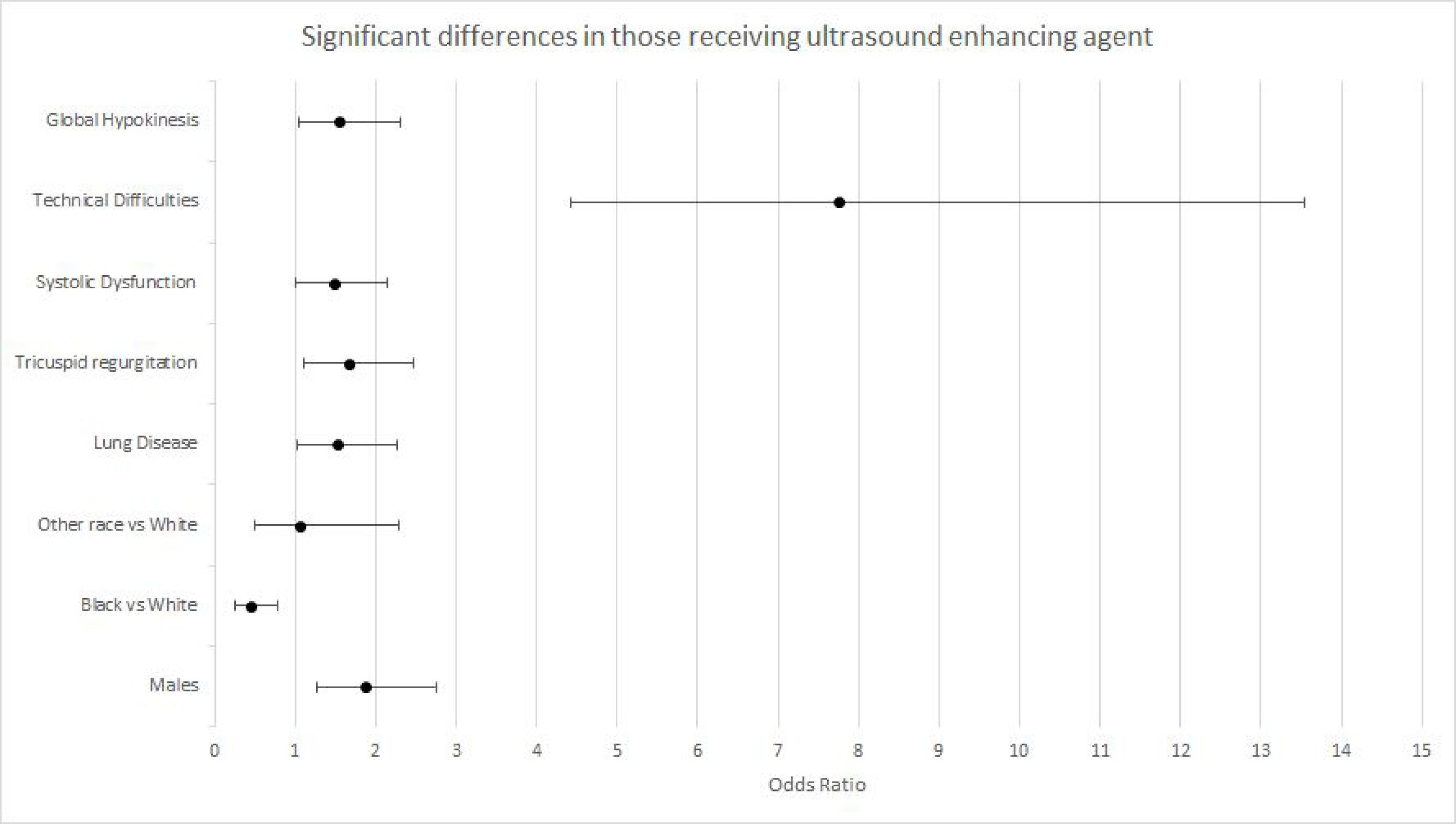
Receiver operating characteristic (ROC) curve for regional wall motion abnormalities (RWMA) on resting transthoracic echocardiography (TTE) predicting coronary artery disease (CAD) adjusting for confounding variables identified during multivariate logistic regression is shown. The area under the curve (AUC) was found to be 0.7136.

### Significant differences in patients with CAD with and without RWMA

As shown in Figure 3, patients with CAD and RWMA were more likely to have a prior history of cancer (OR = 2.35, 95 % CI: 1.2 – 4.6, p = 0.01), systolic dysfunction (OR = 3.37, 95 % CI: 1.9 – 6.0, p < 0.0001), diastolic dysfunction (OR = 4.99, 95 % CI: 2.7 – 9.1, p < 0.0001), and have a TTE indication for non-ST-elevation myocardial infarction (NSTEMI, OR = 2.25, 95 % CI: 1.2 – 4.1, p = 0.008) compared to patients with CAD without RWMA. Patients with CAD and RWMA had a lower BMI (28.2 vs 30.8 kg/m^2^, p = 0.006), lower ejection fraction (EF, 41.6% vs 50.8%, p < 0.0001), and were less likely to have a TTE indication for chest pain (OR = 0.380, 95 % CI: 0.16 – 0.91, p = 0.03) compared to patients with CAD without RWMA. There were no significant differences in gender, prior chemotherapy exposure, heart failure, atrial, atrial flutter, ventricular tachycardia, supraventricular tachycardia, heart block, technical difficulties, global hypokinesis, or the use of UEA.

**Figure 3.**
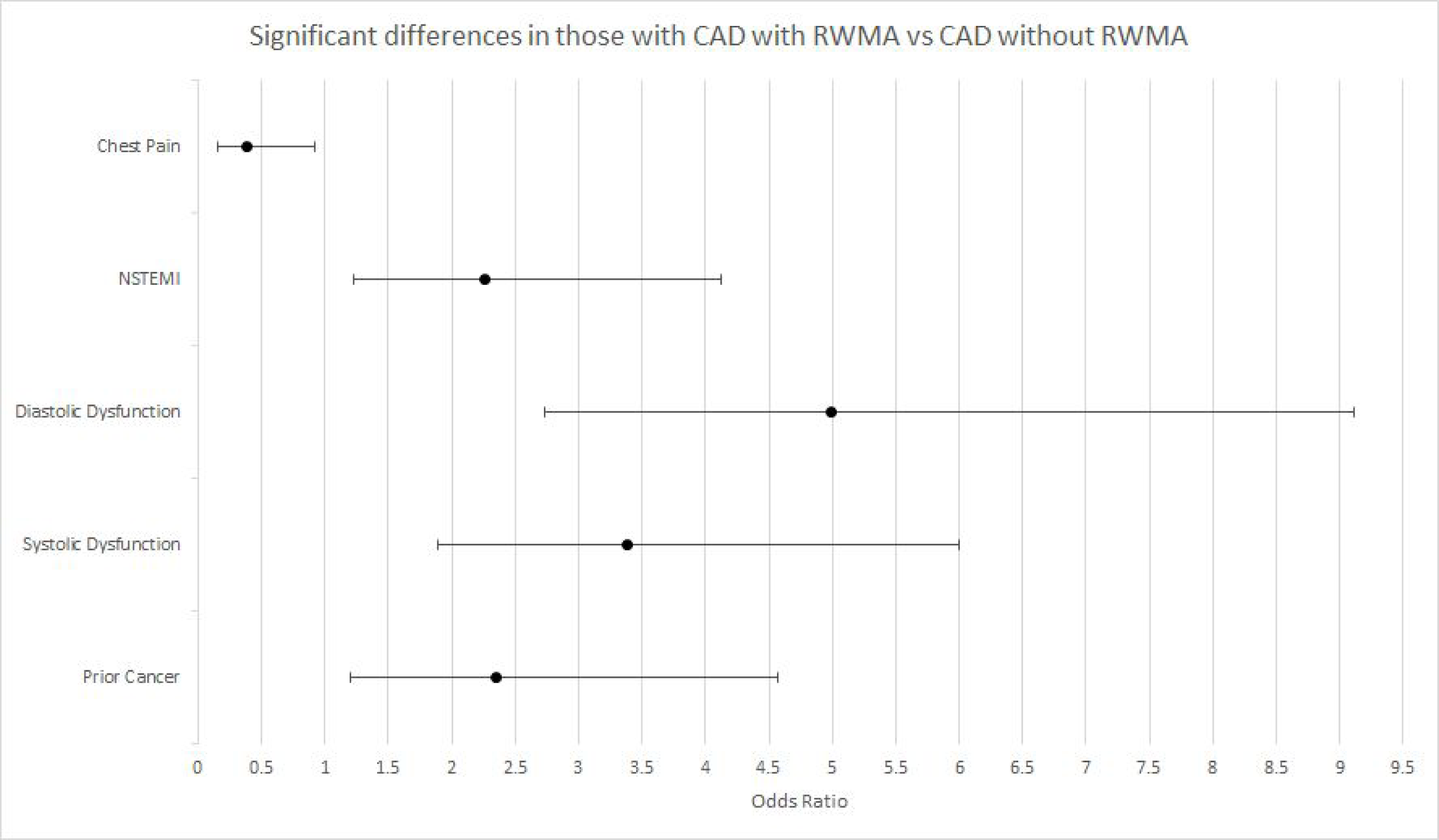
Odds ratio plot of significant, univariate categorical differences between patients with coronary artery disease (CAD) with and without regional wall motion abnormalities (RWMA) on resting transthoracic echocardiography (TTE).

### Impact of Ultrasound Enhancing Agent

Of the 435 included patients, 198 received UEA. As shown in Figure 4, patient who received UEA had a higher BMI (31.6 vs 29.1 kg/m^2^, p = 0.0003) on average and were more likely to be male (OR = 1.88, 95 % CI: 1.3 – 2.8, p = 0.001), have chronic lung disease (OR = 1.52, 95 % CI: 1.02 – 2.3, p = 0.04), have a TTE indication of ST-elevation myocardial infarction (STEMI, p < 0.01), have technical difficulties on TTE (OR = 7.74, 95 % CI: 4.4 – 13.5, p < 0.01), and have global hypokinesis (OR = 1.55, 95 % CI: 1.05 – 2.3, p = 0.03). Patients who received UEA had a lower EF (42.0% vs 46.5%, p = 0.01) on average and were less likely to be black (OR = 0.446, 95 % CI: 0.26 – 0.77, p = 0.004). There were no significant differences in other past medical history, TTE indications, or the presence of RWMA or CAD.

**Figure 4.**
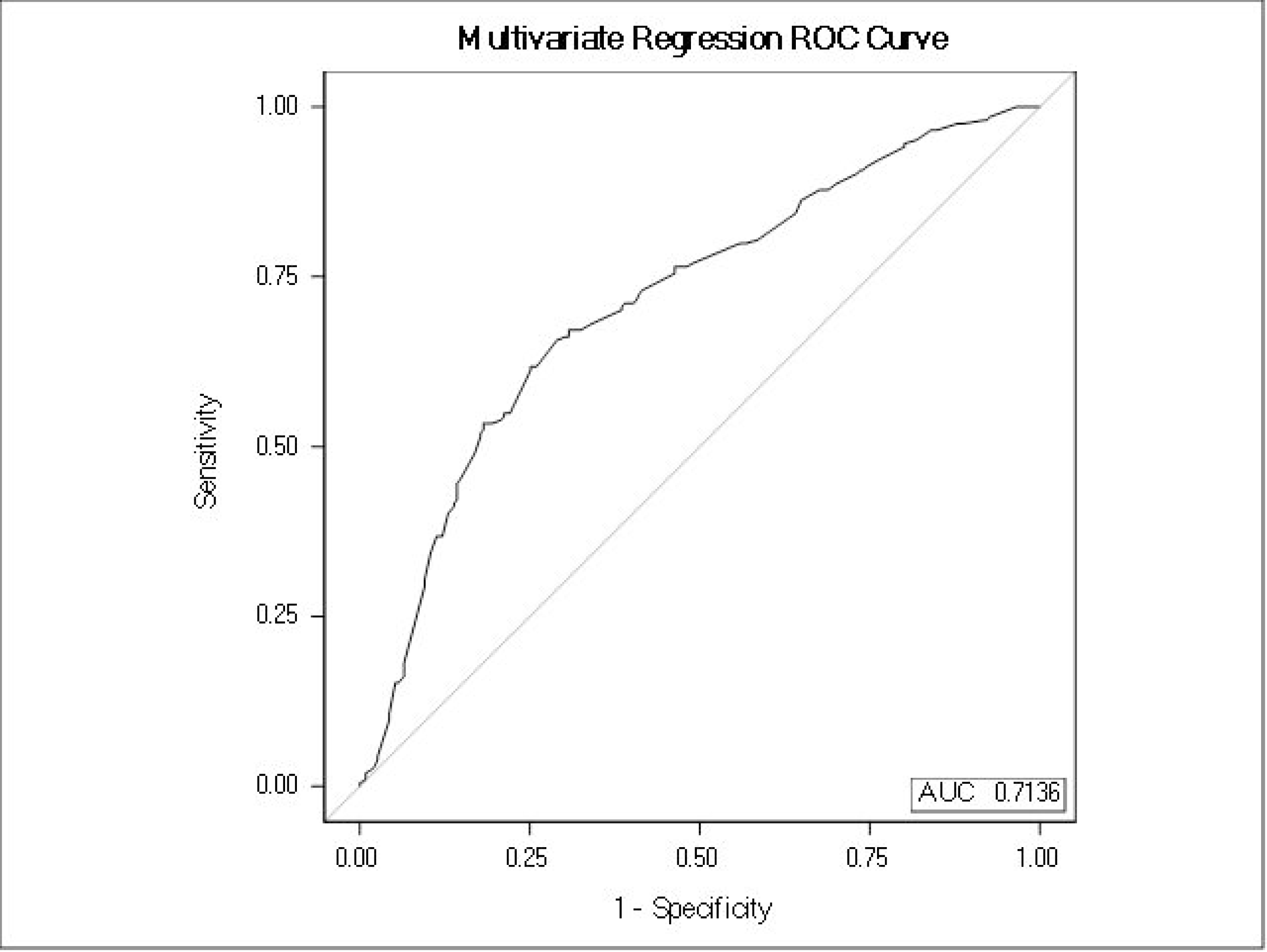
Odds ratio plot of significant, univariate categorical differences between patients who did and did not receive UEA during transthoracic echocardiography.

Of the 198 patients who received UEA, 101 were found to have CAD, 75 were found to have RWMA, and 52 were found to have both CAD and RWMA. Of the 237 patients who did not receive UEA, 103 were found to have CAD, 75 were found to have RWMA, and 49 were found to have both CAD and RWMA. The sensitivity and specificity of RWMA on resting TTE with UEA administration at detecting CAD was 51.5% (95% CI: 48.1 – 54.9%) and 76.3% (95% CI: 61.9 – 90.7%), respectively. The sensitivity and specificity of RWMA on resting TTE without UEA administration at detecting CAD was 47.6% (95% CI: 43.3 – 51.8%) and 80.6% (95% CI = 67.4 – 95.8%), respectively. The sensitivity and specificity of detecting CAD using RWMA was similar between patients who did and did not receive UEA (p = 0.82) (Table 2).

## Discussion

This study demonstrates the ability of RWMA on resting TTE to detect CAD in the adult population in a cardiology practice at a large tertiary care facility. With a sensitivity of 49.5% and negative likelihood ratio of 0.641 (Table 2), TTE has limited ability to rule out significant CAD. Clinicians should be aware of this diagnostic limitation. However, with a specificity of 78.8% and positive likelihood ratio of 2.33, a positive TTE is a stronger indication for catheterization than previously thought.

We demonstrated that RWMA is an insensitive predictor of obstructive CAD in our study’s patient population. Myocardial contractility is not only influenced by perfusion but also mediated by the sympathetic/adrenergic system, integrity of the myocytes and its extracellular matrix, and the differential contributions of the myocardial layers. There may also be some discrepancy between myocardial ischemia and coronary artery stenosis visualized on the coronary angiogram which could introduce bias. With regards to coronary perfusion, the limited sensitivity may be in part due to our cohort also including a small number of outpatient studies where there is a greater prevalence of microvascular dysfunction, usually without detectable wall motion abnormalities. This is in distinct contrast with acute presentations of chest pain where disrupted epicardial coronary flow triggers a wavefront phenomenon that first and foremost affects the myocardial layer with the greatest contribution to contractility, the endocardial layer.(11) In acute presentations of myocardial injury, the mechanism of insult may pose variable responses in wall motion. Not uncommonly there is evidence of clinical and biomarkers of ischemia in the absence of significant coronary artery disease. Myocardial infarction in the absence of obstructive coronary disease (MINOCA) is seen in 6% of acute myocardial infarctions.(12,13) The degree of myocardial depressant and wall motion abnormality may be imperceivable if fibrinolytics are promptly administered. Coupled with the body’s endogenous fibrinolysis, coronary flow may be restored at the time of coronary angiography. In these cases, the evidence of an acute plaque rupture/ulceration may only be appreciated by intravascular ultrasound (IVUS) or optical coherence tomography (OCT). Other causes of MINOCA where RWMA may not be evident include myocarditis in which the mid wall is preferentially affected and Takotsubo cardiomyopathy with a global myocardial depressant phenotype.

The greater specificity for wall motion abnormality in detection of occlusive coronary artery disease can be explained by the ischemic cascade. Myocardial supply-demand mismatch initiates a sequence of events, starting with metabolic disruptions, then perfusion abnormalities, wall motion abnormalities, ECG changes, and angina.(14) Investigative tools such as single-photon emission computed tomography (SPECT) and PET are more sensitive due to detection of ischemia earlier in the cascade (perfusion defects) whereas echocardiography is more specific due to detection of ischemia downstream in the cascade (wall motion abnormalities).

Additionally, we identified several variables which adjust the pre-test probability of finding RWMA in the setting of CAD. Unsurprisingly, increased time between TTE and cardiac catheterization was a negative predictor of RWMA with CAD, likely due to the urgency of catheterization in patients with a positive TTE and suspected CAD. A larger BMI or the presence of systolic or diastolic dysfunction were also negative predictors of RWMA with CAD. These confounding variables may be due to technical limitations, such as inferior image quality of echocardiography in patients with more subcutaneous tissue, and this raises the importance of considering the impact of ultrasound enhancing agent on the sensitivity in more difficult-to-image patient populations.

### Role of UEA in evaluating for CAD

The use of UEA increased sensitivity by 4% and lowered specificity by 4% but these minor changes were statistically insignificant (Table 2). The higher proportion of STEMI patients receiving UEA may be attributed to UEA being used to evaluate for left ventricular thrombus in addition to RWMA. Predictably, the patients who received UEA had a higher average BMI and the TTE reports indicated more frequent technical difficulties. Although UEA does not appear to make a significant difference on the surface, UEA was only administered when necessary to increase the study quality in this difficult-to-image patient population. It is likely that the sensitivity and specificity would be lower in these patients if they had not received UEA and that UEA increased the sensitivity and specificity towards the expected values seen in patients who did not receive UEA. This study should not discourage the use of UEA when necessary but also does not provide clear evidence that UEA should be more widely used when evaluating for RWMA. To further evaluate the effect of UEA on the relationship between RWMA on resting TTE and obstructive CAD, a prospective study evaluating pre- and post-UEA resting TTE interpretations would be required.

### Comparison to previous studies

Upon review of the English cardiovascular literature, to our knowledge this is the largest study with the most inclusive patient population to date to correlate RWMA on TTE to significant CAD. Whereas this study included all patients who received both a TTE and cardiac catheterization coronary angiography for any reason, previous studies focused on patients with chest pain, patients status post return of spontaneous circulation (ROSC) following cardiac arrest (CA) from a cardiac etiology, patients suspected of having MI, and patients with left ventricular dysfunction on TTE (Table 3).(5–8,15,16) We report a lower sensitivity from the previous range reported across various studies of 63.6% - 93.1%.(5–8,16) Compared to Sasaki et al., we report a higher sensitivity and specificity.(15) We report a higher specificity than Kim et al., Sabia et al., and Medina et al.(5,7,16) We report a lower specificity than Lee et al. and Jacobsen et al.(6,8) This comparison is summarized in Table 3.

**Table 3.** Comparison of this study to previous studies investigating the diagnostic reliability of regional wall motion abnormalities (RWMA) on resting transthoracic echocardiography (TTE) to predict significant coronary artery disease (CAD, ≥ 70% vessel occlusion).

| Study | This Study | Sabia et al.<sup>5</sup> | Jacobsen et al.<sup>8</sup> | Sasaki et al.<sup>15</sup> | Lee et al.<sup>9</sup> | Kim et al.<sup>7</sup> | Medina et al.<sup>16</sup> |
| --- | --- | --- | --- | --- | --- | --- | --- |
| Year | 2023 | 1991 | 2022 | 1986 | 2015 | 2021 | 1985 |
| Study Type | Retrospective | Prospective | Prospective | Prospective | Prospective | Retrospective | Retrospective |
| Study Population | All adults | Suspected MI | First response, Chest Pain | Chest Pain | Chest Pain | s/p ROSC from CA | RWMA or global hypokinesis on echo |
| Total, n | 435 | 169 | 240 | 46 | 69 | 145 | 103 |
| CAD, n | 204 | 29 | 22 | 8 | 25 | 76 | 87 |
| RWMA, n | 150 | 87 | 33 | 18 | 21 | 105 | 83 |
| CAD + RWMA, n | 101 | 27 | 14 | 3 | 16 | 68 | 77 |
| Sensitivity | 49.5% | 93.1% | 63.6% | 37.5% | 64.0% | 89.5% | 88.5% |
| Specificity | 78.8% | 57.1% | 91.2% | 60.5% | 88.6% | 46.4% | 62.5% |

The variability of results in the literature may reflect differences in the study methodology or patient cohorts. Additionally, differences in pre-test probability of CAD likely contribute to the diagnostic variability, given the range of populations and CAD prevalence across the studies.

### Limitations of this study

The retrospective design of this study introduces inherent limitations, such as difficulty with assigning causation between wall motion abnormalities and obstructive coronary disease. Confounding is typically more prevalent in retrospective designs; for example, the presence of myocarditis and/or coronary recanalization may impact degree of wall motion abnormality but were not accounted for in the absence of IVUS/OCT or cardiac magnetic resonance imaging (MRI). Inter-reader differences among echocardiographers and angiographers may reflect varying sensitivities, however, our study reflects a real-world large cardiology practice with complex coronary pathologies.

Although ECG and cardiac biomarkers are valuable tools in the workup of a patient with ACS, this study did not attempt to correlate ECG or biomarker findings with RWMA. Patients with RWMA were more likely to have a TTE indication for NSTEMI and we expect a positive correlation between ECG and biomarker findings suggestive of MI with RWMA, given the correlation between RWMA and CAD found here.

This study included all adult patients, including those with technical difficulties and adequate quality TTEs, to better indicate the true diagnostic usefulness of TTE applied to a broad patient population. The diagnostic reliability of TTE at detecting CAD may be highly dependent upon patient selection, and stratifying groups to evaluate which populations TTE is most reliable for may be an area for future exploration.

## Conclusion

TTE is a valuable, non-invasive method for evaluating patients with suspected CAD, especially in patients suspected of having MI or ACS but with inconsistent history, ECG, and/or biomarkers. Our study found a sensitivity of 49.5%, specificity of 78.8%, positive likelihood ratio of 2.33, and negative likelihood ratio of 0.641 for the presence of RWMA on resting TTE to predict significant CAD in an adult population in both outpatient and inpatient settings The lower sensitivity is due to the complex pathobiology of myocardial contractility and the higher specificity is due to the wall motion abnormality being at the latter part of the ischemic cascade. The use of ultrasound enhancing agent did not enhance nor detract from the sensitivity or specificity but may be necessary in difficult-to-image patients. Clinicians should be aware of these diagnostic limitations, particularly the relatively low sensitivity, as they apply the use of TTE to patients with suspected CAD.

## Data Availability

Data are available at request

## References

1. Duggan JP, Peters AS, Trachiotis GD, Antevil JL. Epidemiology of Coronary Artery Disease. Surg Clin North Am 2022;102:499–516.

2. Sirtori CR, Labombarda F, Castelnuovo S, Perry R. The use of echocardiography for the non-invasive evaluation of coronary artery disease. Ann Med 2017;49:134–141.

3. American College of Cardiology Foundation Appropriate Use Criteria Task F, American Society of E, American Heart A et al. ACCF/ASE/AHA/ASNC/HFSA/HRS/SCAI/SCCM/SCCT/SCMR 2011 Appropriate Use Criteria for Echocardiography. A Report of the American College of Cardiology Foundation Appropriate Use Criteria Task Force, American Society of Echocardiography, American Heart Association, American Society of Nuclear Cardiology, Heart Failure Society of America, Heart Rhythm Society, Society for Cardiovascular Angiography and Interventions, Society of Critical Care Medicine, Society of Cardiovascular Computed Tomography, and Society for Cardiovascular Magnetic Resonance Endorsed by the American College of Chest Physicians. J Am Coll Cardiol 2011;57:1126–66.

4. Beller GA. Myocardial perfusion imaging for detection of silent myocardial ischemia. Am J Cardiol 1988;61:22F - 26F.

5. Sabia P, Afrookteh A, Touchstone DA, Keller MW, Esquivel L, Kaul S. Value of regional wall motion abnormality in the emergency room diagnosis of acute myocardial infarction: a prospective study using two-dimensional echocardiography. Circulation 1991;84:I-85 - I-92.

6. Lee M, Chang SA, Cho EJ et al. Role of strain values using automated function imaging on transthoracic echocardiography for the assessment of acute chest pain in emergency department. Int J Cardiovasc Imaging 2015;31:547–56.

7. Kim J, Cho YS, Lee BK et al. Diagnostic value of transthoracic echocardiography compared to electrocardiogram in predicting coronary artery stenosis among patients after cardiac arrest. Am J Emerg Med 2021;46:97–101.

8. Jacobsen L, Grenne B, Olsen RB, Jortveit J. Feasibility of prehospital identification of non-ST-elevation myocardial infarction by ECG, troponin and echocardiography. Emerg Med J 2022;39:679–684.

9. Lee SE, Uhm JS, Kim JY, Pak HN, Lee MH, Joung B. Combined ECG, Echocardiographic, and Biomarker Criteria for Diagnosing Acute Myocardial Infarction in Out-of-Hospital Cardiac Arrest Patients. Yonsei Med J 2015;56:887–94.

10. Mitchell C, Rahko PS, Blauwet LA et al. Guidelines for Performing a Comprehensive Transthoracic Echocardiographic Examination in Adults: Recommendations from the American Society of Echocardiography. J Am Soc Echocardiogr 2019;32:1–64.

11. Kim RJ, Wu E, Rafael A et al. The Use of Contrast-Enhanced Magnetic Resonance Imaging to Identify Reversible Myocardial Dysfunction. New England Journal of Medicine 2000;343:1445–1453.

12. Pasupathy S, Air T, Dreyer RP, Tavella R, Beltrame JF. Systematic review of patients presenting with suspected myocardial infarction and nonobstructive coronary arteries. Circulation 2015;131:861–70.

13. Tamis-Holland JE, Jneid H, Reynolds HR et al. Contemporary Diagnosis and Management of Patients With Myocardial Infarction in the Absence of Obstructive Coronary Artery Disease: A Scientific Statement From the American Heart Association. Circulation 2019;139:e891–e908.

14. Maznyczka A, Sen S, Cook C, Francis DP. The ischaemic constellation: an alternative to the ischaemic cascade-implications for the validation of new ischaemic tests. Open Heart 2015;2:e000178.

15. Sasaki H, Charuzi Y, Beeder C, Sugiki Y, Lew AS. Utility of echocardiography for the early assessment of patients with nondiagnostic chest pain. American Heart Journal 1986;112:494–497.

16. Medina R, Panidis IP, Morganroth J, Kotler MN, Mintz GS. The value of echocardiographic regional wall motion abnormalities in detecting coronary artery disease in patients with or without a dilated left ventricle. American Heart Journal 1985;109:799–803.

